# Heterogeneity in Early Postpartum Metabolic Profiles Among Women with GDM Who Progressed to Type 2 Diabetes During 10-Year Follow-Up: The SWIFT Study

**DOI:** 10.1101/2023.06.13.23291346

**Authors:** Saifur R Khan, Hannes Rost, Brian Cox, Babak Razani, Stacey Alexeeff, Michael B. Wheeler, Erica P. Gunderson

## Abstract

GDM is a strong risk factor for progression to T2D after pregnancy. Although both GDM and T2D exhibit heterogeneity, the link between the distinct heterogeneity of GDM and incident T2D has not been established. Herein, we evaluate early postpartum profiles of women with recent GDM who later developed incident T2D using a soft clustering method, followed by the integration of both clinical phenotypic variables and metabolomics to characterize these heterogeneous clusters/groups clinically and their molecular mechanisms. We identified three clusters based on two indices of glucose homeostasis at 6-9 weeks postpartum – HOMA-IR and HOMA-B among women who developed incident T2D during the 12-year follow-up. The clusters were classified as follows: pancreatic beta-cell dysfunction group (cluster-1), insulin resistant group (cluster-3), and a combination of both phenomena (cluster-2) comprising the majority of T2D. We also identified postnatal blood test parameters to distinguish the three clusters for clinical testing. Moreover, we compared these three clusters in their metabolomics profiles at the early stage of the disease to identify the mechanistic insights. A significantly higher concentration of a metabolite at the early stage of a T2D cluster than other clusters indicates its essentiality for the particular disease character. As such, the early-stage characters of T2D cluster-1 pathology include a higher concentration of sphingolipids, acyl-alkyl phosphatidylcholines, lysophosphatidylcholines, and glycine, indicating their essentiality for pancreatic beta-cell function. In contrast, the early-stage characteristics of T2D cluster-3 pathology include a higher concentration of diacyl phosphatidylcholines, acyl-carnitines, isoleucine, and glutamate, indicating their essentiality for insulin actions. Notably, all these biomolecules are found in the T2D cluster-2 with mediocre concentrations, indicating a true nature of a mixed group. In conclusion, we have deconstructed incident T2D heterogeneity and identified three clusters with their clinical testing procedures and molecular mechanisms. This information will aid in adopting proper interventions using a precision medicine approach.

## INTRODUCTION

Type 2 diabetes (T2D) is a chronic disease characterized by pancreatic beta-cell dysfunction and insulin insensitivity. Although the risk of T2D onset is tied to genetic predisposition, as can be gleaned through family history, it depends on a summation of environmental factors and lifestyle behaviors [1, 2]. These intricacies of T2D are further complicated by the heterogeneity of the disease, which, in part, can be explained by the relative contributions of insulin resistance and beta-cell dysfunction, among other factors [3]. Understanding this heterogeneity is essential as treatment regimens move towards a personalized medicine approach [4, 5].

A method for deconstructing T2D heterogeneity is cluster analysis [5], commonly using biochemical variables, clinical characteristics, and genetics. For example, Ahlqvist and colleagues used a hard-clustering approach (i.e., k-means and hierarchical clustering) on six variables [i.e., glutamate decarboxylase antibodies, age at diagnosis, body mass index (BMI), HbA1c, and a homeostatic model assessment estimating β-cell function and insulin resistance] of the ‘Swedish All New Diabetics in Scania cohort’ [6]. This cohort included only the diabetes population, with approximately 80% classified as T2D. They identified five clusters based on the degree of insulin deficiencies and insulin resistance, then compared them for disease progression, treatment, and development of diabetic microvascular complications. For example, cluster-2, which belongs to insulin-deficient groups, exhibited the highest risk of retinopathy, whereas cluster-3 showed the highest degree of insulin resistance and possessed the highest risk of diabetic kidneys. Li and colleagues identified three subgroups of T2D patients using topology-based patient-patient networks (an unsupervised soft network model) on the electronic medical records and genomic profiles [7]. Recently, Udler and colleagues explored genomic profiles and traits of T2D heterogeneous groups using a soft clustering method (i.e., Bayesian nonnegative matrix factorization clustering) and identified five genetic variant-trait associated clusters using only T2D population of Caucasian ancestry from the Metabolic Syndrome in Men Study (METSIM), the Diabetes Genes in Founder Populations (Ashkenazi) study, the Partners HealthCare Biobank, and the UK Biobank [8]. This analysis was further extended to estimate genetic risk scores for distinct clinical outcomes.

Gestational diabetes mellitus (GDM) is a transient form of diabetes that develops during mid-gestation of pregnancy. Although the vast majority of women with GDM return to pre-pregnancy glucose tolerance (i.e., euglycemia or prediabetes) postpartum, about 30% of women with GDM are classified with prediabetes in the postpartum period and another 5% or more with type 2 diabetes depending on the population [9]. Overall, a history of GDM is associated with a 4-to 10-fold higher risk of future T2D [10] although few studies have ruled out pre-pregnancy type 2 diabetes within the GDM diagnosis[11]. As such, GDM is a harbinger of future T2D risk, and offers an opportunity for early prevention by identifying and differentiating early-stage biomarkers preceding clinical pathophysiological for prediabetes and diagnosis of T2D [12-16]. Powe and colleagues devised a hard clustering method (i.e., calculation of 25^th^ percentile of normal glucose tolerance) on the Genetics of Glucose regulation in the Gestation and Growth (Gen3G) cohort. They defined three subgroups of women with GDM (primarily beta-cell dysfunction, primarily insulin resistance, and mixed) [17]. Liu and colleagues adopted this hard-clustering method and identified the same GDM groups in a cohort of Shanghai Jiao-Tong University Affiliated Sixth People’s Hospital [18]. However, it is unknown whether GDM heterogeneity contributes to future T2D heterogeneity. As such, it is important to decipher the heterogeneity of the early postpartum profiles among women with recent GDM who will later progress to T2D. Additionally, the application of a soft clustering method (i.e., allowing data of a participant to be part of multiple groups based on data quality and finding out the participant’s true character) allows researchers to minimize the biases of the results. Furthermore, the integration of ‘omics, and the addition of clinical variables, could identify the molecular mechanisms of disease pathophysiology. In this case, metabolomics is expected to be highly informative due to the prominent influence of environmental factors and lifestyle behaviors on these diseases (i.e., GDM and T2D).

Motivated to fill these knowledge gaps, we applied a soft clustering method to the study baseline (6-9 weeks postpartum) indices of fasting plasma glucose and insulin among subsequent incident T2D cases (n=226, 22% of 1010 without T2D at baseline) identified prospectively during 10 years of follow up of the Study of Women, Infant Feeding and Type 2 Diabetes after GDM (SWIFT Study). The SWIFT study is a prospective GDM research cohort that enrolled 1,035 women with GDM from 2008-2011 from Kaiser Permanente Northern California into three in-person research examinations with 2-h 75 g OGTTs at 6-9 weeks postpartum and annually for two years post-baseline, and supplemented by clinical diagnoses of diabetes from electronic health records up to 10 years [19, 20]. We deconstruct their heterogeneity, followed by integration of both clinical research phenotypic variables and metabolomics to characterize these heterogeneous clusters/groups clinically and molecular levels.

### Identification of incident case clusters

We have identified three distinct incident case clusters within the total incident cases of T2D during follow up among 1,010 of the 1,033 women with recent GDM without T2D at 6-9 weeks postpartum. A total of 226 women with GDM developed incident T2D within 10-12 years post-delivery (October 2020). The analysis includes 225 incident T2D cases after exclusion of 1 woman with T2D missing fasting insulin at baseline. Using fasting plasma samples for the 2-h 75 g research OGTTs (**Figure-1, 2A**) we calculated HOMA-B and HOMA-IR indices from the fasting glucose and insulin research measurements (Northwest Lipid Research Laboratories, University of Washington, Seattle, WA) at study baseline (6-9 weeks of postpartum) and identify three incident T2D clusters. Cluster-1 consists of 81 incident T2D cases, Cluster-2 consists of 120 incident T2D cases, and Cluster-3 includes 24 incident T2D cases. Each cluster is differentiated by the HOMA-B and HOMA-IR values (**Figure-2B, 2C**). Cluster-1 possesses comparatively the worst pancreatic beta-cell function [i.e., HOMA-B, Median (IQR): 161.3 (126.8-202.4)] but the best insulin sensitivity [i.e., HOMA-IR, Median (IQR): 3.82 (2.97-4.59)]. As such, we term this cluster-1 as the ‘severe pancreatic beta-cell dysfunction’ group. The cluster-3 possesses comparatively the best pancreatic beta-cell function [i.e., HOMA-B, Median (IQR): 630.47 (454.8-724.9)] but the worst insulin sensitivity [i.e., HOMA-IR, Median (IQR): 16.83 (11.99-23.27)]. As such, we term this cluster-3, the ‘severe insulin resistance’ group. Cluster-2 possesses a comparatively moderate level of both pancreatic beta-cell function [i.e., HOMA-B, Median (IQR): 307.24 (263.97-389.5)] and insulin sensitivity [i.e., HOMA-IR, Median (IQR): 8.63 (6.70-10.50)]. Cluster-3 is therefore termed “Intermediate pancreatic beta-cell dysfunction and insulin-resistance group.

**Figure 1:**
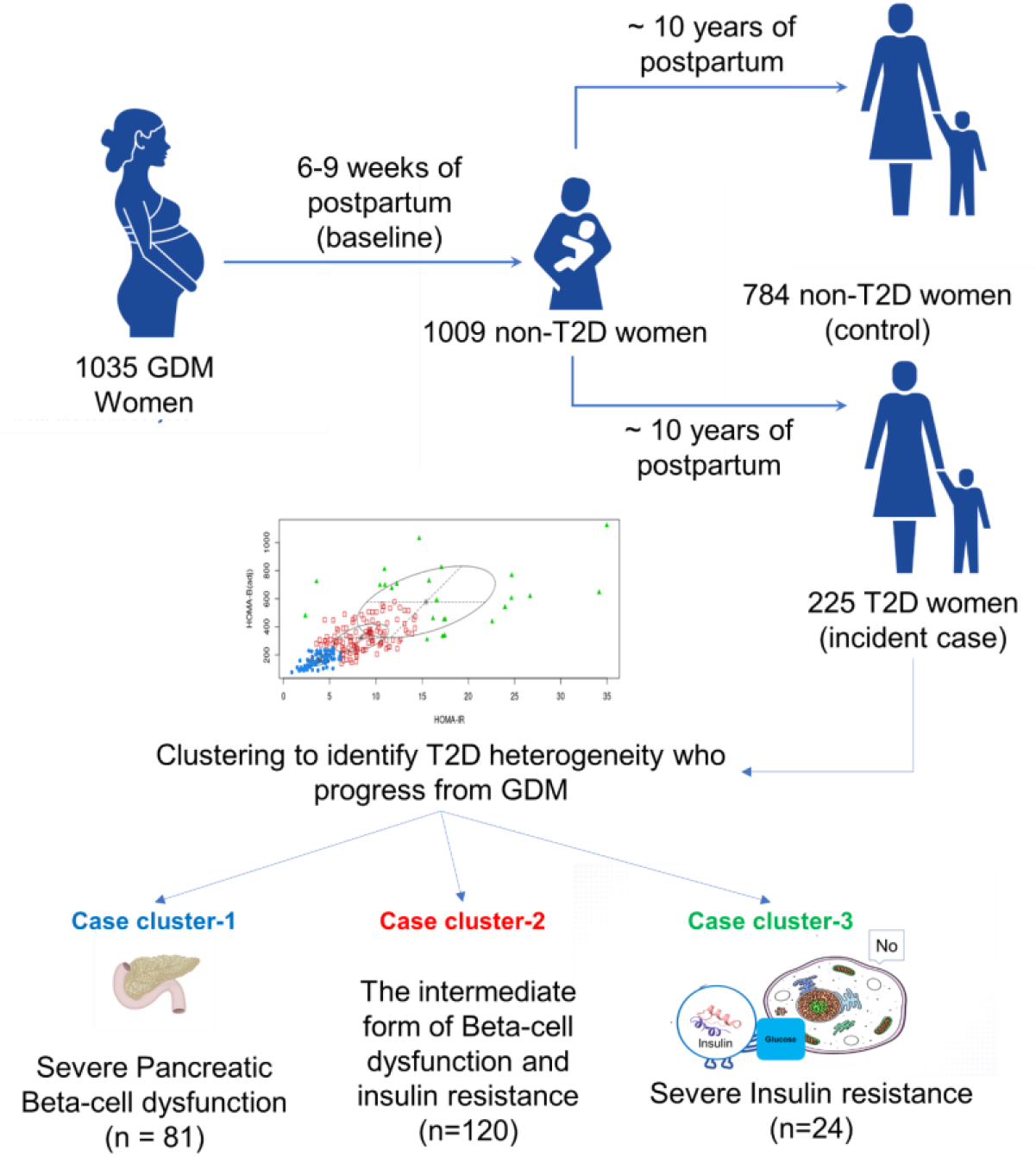
The flow diagram of the study design. Two baseline clinical variables (i.e., HOMA-IR and HOMA-B) of 225 incident cases (who developed T2D within ten years of postpartum) were subjected to unsupervised model-based clustering using the *mclust* algorithm to identify the heterogeneous groups. Later, these groups were defined using their baseline other clinical variables, T2D onset time, and metabolomics data.

**Figure 2:**
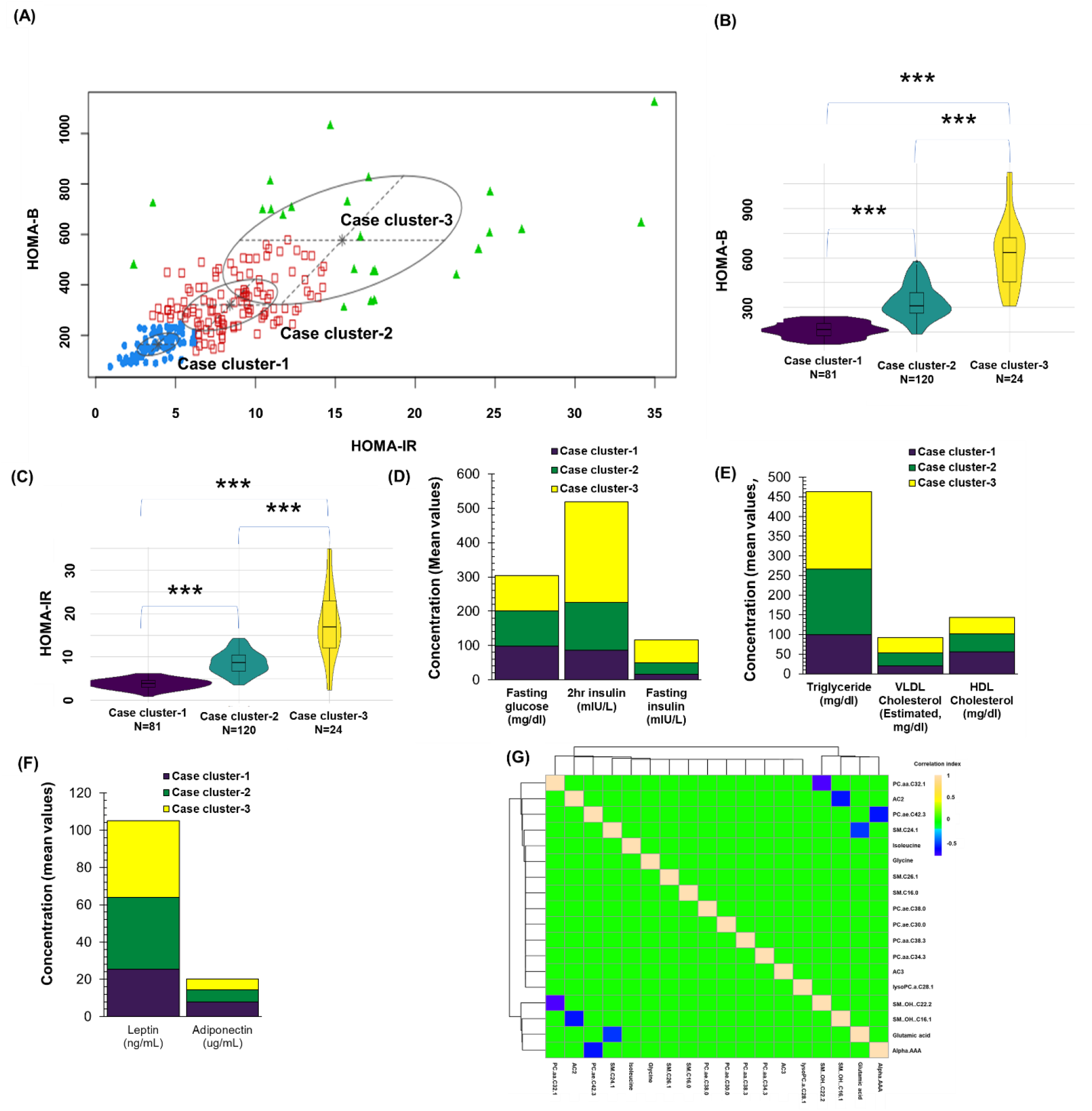
Heterogeneous group identification and characterization. (A) The *mclust* algorithm identifies three clusters within the incident cases based on HOMA-B and HOMA-IR. (B) The violin box plots of HOMA-B values between these three clusters show distinct and significant separation. (C) The violin box plots of HOMA-IR values between these three clusters show distinct and significant separation. (D-F) A comparison of some major clinical variables (i.e., fasting glucose, 2h-insulin, fasting insulin, triglyceride, VLDL cholesterol, HDL cholesterol, leptin, and adiponectin) between these clusters. (G) ANOVA identified 18 significant metabolites at the baseline between these clusters. A correlation heatmap for the 18 significant metabolites with a correlation cut-off ± 0.5.

### Socio-demographic and phenotypic characteristics of case clusters

The 1,033 women with GDM (recruited during pregnancy) within the SWIFT prospective follow up cohort comprise highly diverse racial and ethnic groups (75% minority: Asian, Hispanic, Black, 23% White, 2% other). The racial and ethnic distribution of incident T2D populations in different clusters are not significantly different. In the case of mean age, the incident T2D population of cluster-1 (i.e., 35.50 ± 0.48) is significantly older than both cluster-2 (i.e., 33.47 ± 0.47) and cluster-3 (i.e., 30.87 ± 1.22) (*P* = 0.0001). However, the opposite trend is observable in pre-pregnancy BMI and study baseline (6-9 weeks of postpartum) BMI, weight, and waist circumference. The mean pre-pregnancy BMI of cluster-1 (i.e., 30.45 ± 0.79) is significantly lower than both cluster-2 (i.e., 35.39 ± 0.77) and cluster-3 (i.e., 36.09 ± 1.40) (*P* < 0.0001). Similarly, cluster-1 women exhibit significantly lower baseline BMI, weight, and waist circumference than those of both cluster-2 and cluster-3. This suggests that pancreatic beta-cell dysfunction (i.e., cluster-1) may be age-dependent, whereas the insulin resistance (i.e., cluster-3) is obesity-dependent. The median time to diabetes onset was very similar for cluster-1 and cluster-2; about 35 and 41 months, respectively, but there was a trend for cluster-3 to exhibit a shorter time to diabetes onset of 22 months (*P* = 0.21). These three clusters were not statistically different in terms of parity, GDM prenatal type of treatment, gestational age of GDM diagnosis (*P* = 0.07), hypertension history, and family history of diabetes (**Table-1**).

**Table 1:**
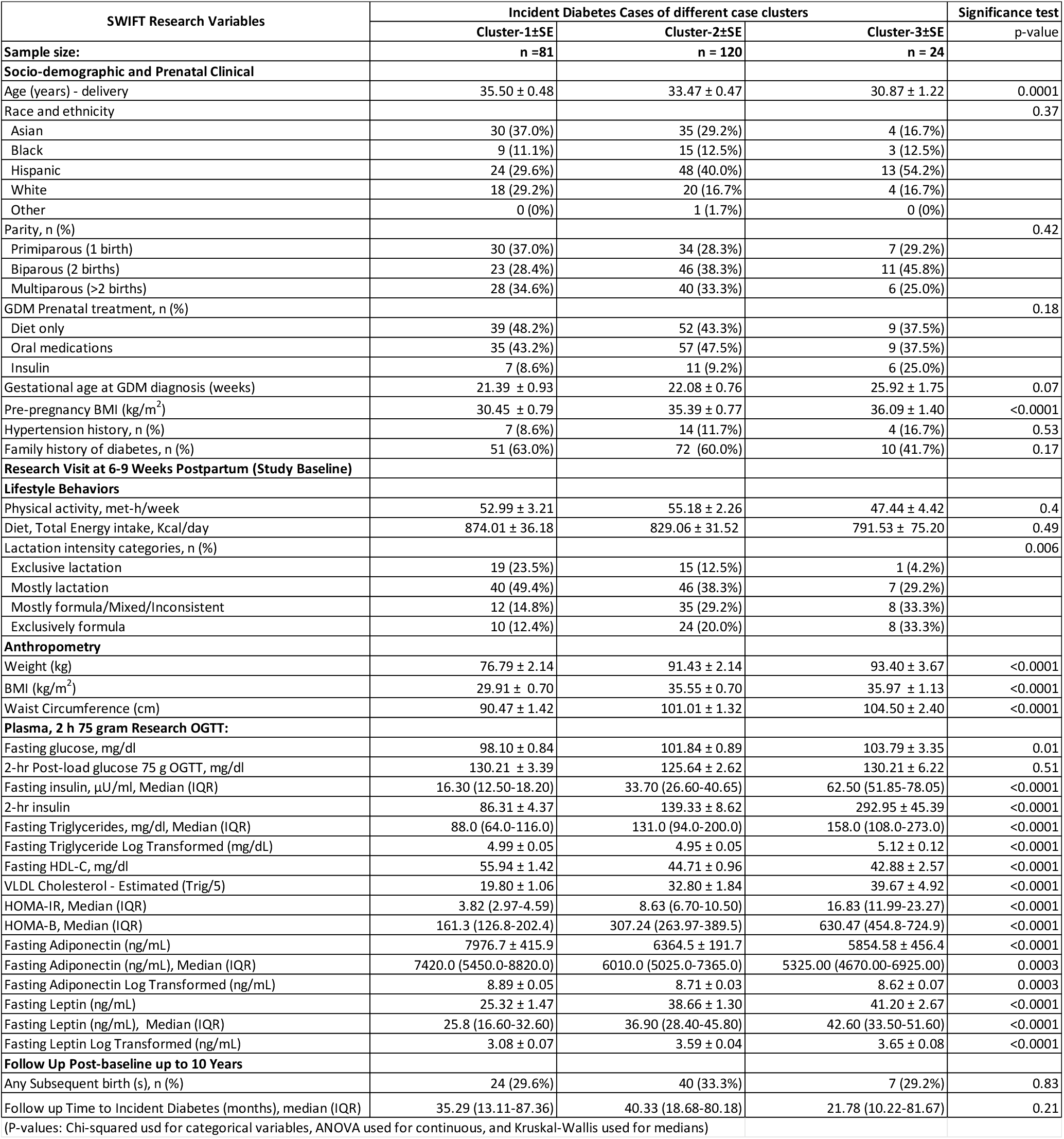
The characteristics of three case clusters. Table X. title of baseline characteristics for incident cases of Diabetes at 10-year follow up

### Research variables of case clusters

The three incident T2D clusters were significantly different based on numerous research parameters including fasting plasma glucose and insulin, 2h post-load plasma insulin, fasting plasma triglycerides, HDL-cholesterol, VLDL-cholesterol, and adiponectin and leptin (**Figure 2D-2F, Table-1**). In brief, the requirement of insulin is increased with the degree of insulin resistance (i.e., cluster-1 < cluster-2 < cluster-3). Although cluster-3 requires the highest level of insulin, this cluster exhibits significantly worsening of fasting glucose (**Table-1**). Fasting triglycerides and VLDL-cholesterols were also increased at 6-9 weeks postpartum among the incident T2D cases with greater severity of their heightened insulin resistance. For instance, cluster-3 possesses significantly higher levels of triglycerides and VLDL-cholesterol than those of cluster-2 and cluster-1 (**Table-1**). The opposite trend (with statistical significance) can be observed for fasting HDL-cholesterol and fasting adiponectin concentrations (**Table-1)**. These two biological molecules are well-known for their positive roles in breakdown of triglycerides and in reducing insulin resistance. With respect to the anorexic adipokine Leptin, cluster-3 and -2 possessed significantly higher levels than cluster-1 (**Table-1)**. As expected, these two clusters have high proportions of clinically obese women (i.e., BMI ≥ 30) (**Table-1)**. It suggests that there is a leptin resistance in cluster-2 and -3.

### Effects of breast-feeding in the T2D progression

Breast-feeding which has a well-known association to the protection against T2D progression, is correlated to suppression of lipogenesis in the liver [22]. However, higher GDM severity, (i.e., maternal hyperglycemia requiring insulin treatment) and pre-pregnancy obesity have been clinically associated with delayed onset of lactogenesis II (i.e., the onset of copious milk production) which can alter the breast-feeding behavior [23].). We measured lactation intensity at 6-9 weeks postpartum associated with cluster phenotypes. For cluster-1, 72.9% of the incident T2D cases (i.e., severe pancreatic beta-cell dysfunction group) were found to be in the fully/exclusive or mostly lactation groups at baseline, as compared with 50.8% of cluster-2 and 33.4% of cluster-3 (**Table-1**). Lactation was significantly higher in cluster-1 than other clusters. Since there was no significant difference in GDM treatment among the lactation intensity groups at baseline, the severity of GDM did not determine lactation behavior among these clusters. As stated above, cluster-2 and -3 exhibited significantly higher BMI, weight, and waist circumference than cluster-1 (**Table-1)**. With respect to race/ethnicity, clusters 2 and 3 included higher proportions of Hispanic, and lower proportions of Asian groups, consistent with the prominence of clinically obese cases (i.e., BMI ≥ 30). Altogether, lower lactation of cluster 2 and 3 could be attributed to obesity related delayed onset of lactogenesis II, many other factors determine lactation practices such as lower breastfeeding support for obese mothers, social and economic barriers, clinical outcomes, and cultural practices. Additionally, pancreatic beta cell dysfunction with higher intensity lactation may be a contributing factor in the lower fasting insulin. Thus, cluster 1 profile may be a result of higher intensity lactation, and cluster 2 and 3 may result from increased insulin secretion at baseline related to lower lactation.

### The molecular characteristic of the early-stage T2D pathophysiology between case clusters

We have utilized baseline (6-9 weeks postpartum) blood plasma metabolomics data to distinguish the case cluster-specific early-stage pathophysiology at molecular levels. These metabolomics data consist of 188 metabolites from different chemical groups such as amino acids, fatty acids, phospholipids, sphingolipids, and carnitines. A total of eighteen metabolites were found significantly different between case clusters under a corrected *P*-value for multiple testing using Benjamini–Hochberg method and used FDR cut-off < 0.05 (**Figure-2H**). These metabolites belong to one of following chemical groups: phospholipids, sphingolipids, carnitines, and amino acids.

We have found seven phospholipids are significantly altered between the case clusters. These are: three phosphatidylcholine diacyl molecules (i.e., PC aa C32:1, PC aa C34:3, PC aa C38:3), three phosphatidylcholine acyl-alkyl molecules/plasmalogens (PC ae C30:0, PC ae C38:0, and PC ae C42:3), and one lysophosphatidylcholine acyl molecule (i.e., lysoPC a 28:1). We observe the blood concentration trend of significantly altered phosphatidylcholine diacyl molecules increase from case cluster-1 towards case cluster-3. However, an opposite trend can be observed in the case of significantly altered phosphatidylcholine acyl-alkyls/plasmalogens and lysophosphatidylcholine acyls (**Figure 3**). Additionally, we have found five sphingolipids are significantly altered between the case clusters. These are SM C16:0, SM(OH)C16:1, SM(OH)C22:2, SMC24:1, and SMC26:1. We observed that the plasma concentration trend of significantly altered sphingolipids decline from case cluster-1 towards cluster-3 (**Figure 4**). Further, the three case clusters were significantly different in blood concentrations of two acetylcarnitines (i.e., AC2 and AC3) and four amino acids (i.e., glutamate, glycine, alpha.AAA [α-aminoadipic acid], and isoleucine) (**Figure 5**). We observed an upward concentration trend in the significantly altered acetylcarnitines and all amino acids (except glycine) from case cluster-1 towards cluster-3. Glycine was the exception exhibiting an opposite trend being highest in cluster 1.

**Figure 3:**
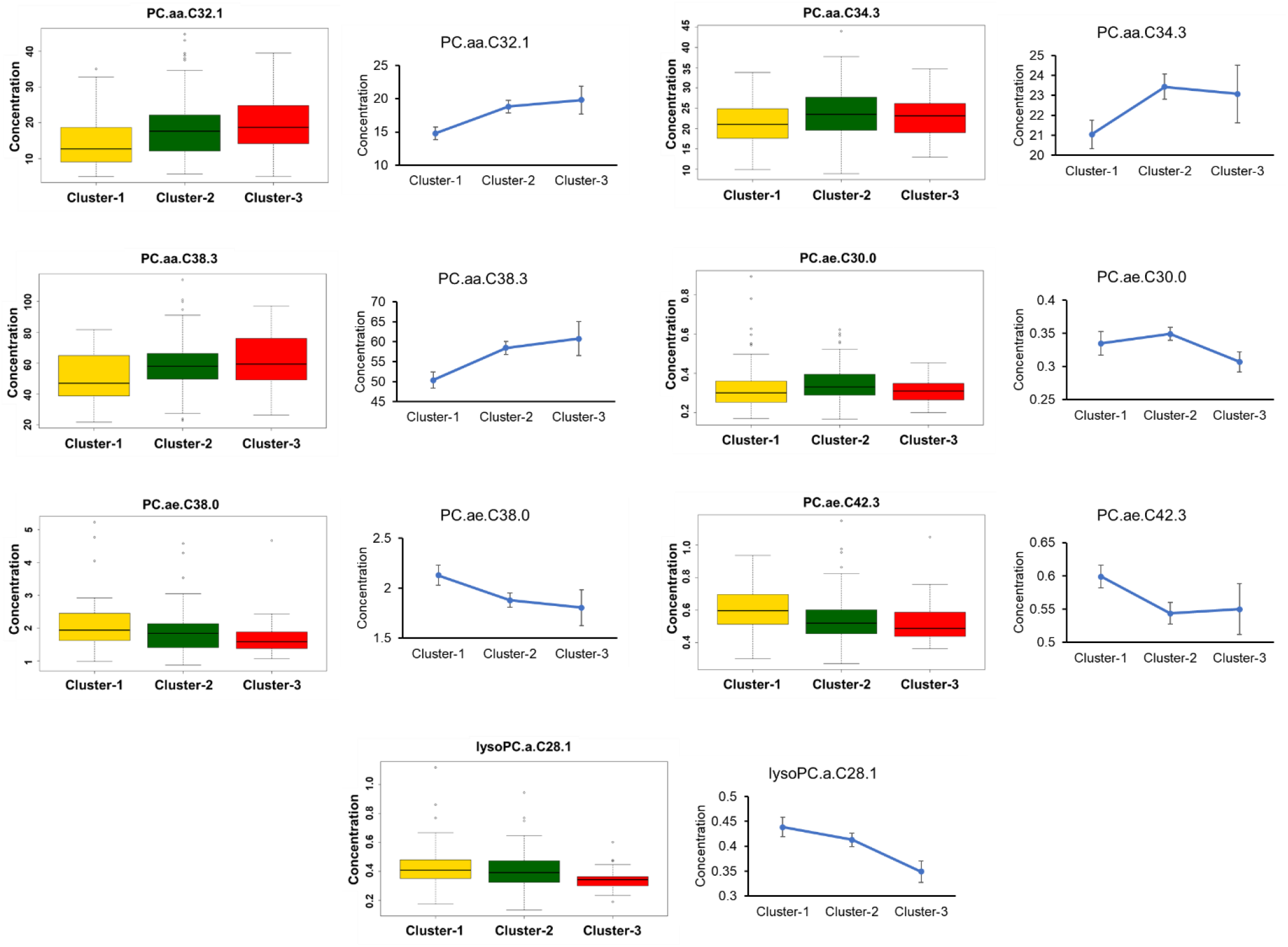
The significantly altered phospholipid species between clusters. A box plot and adjacent line plot were used here to display the variations between the cluster for each significant phospholipid.

**Figure 4:**
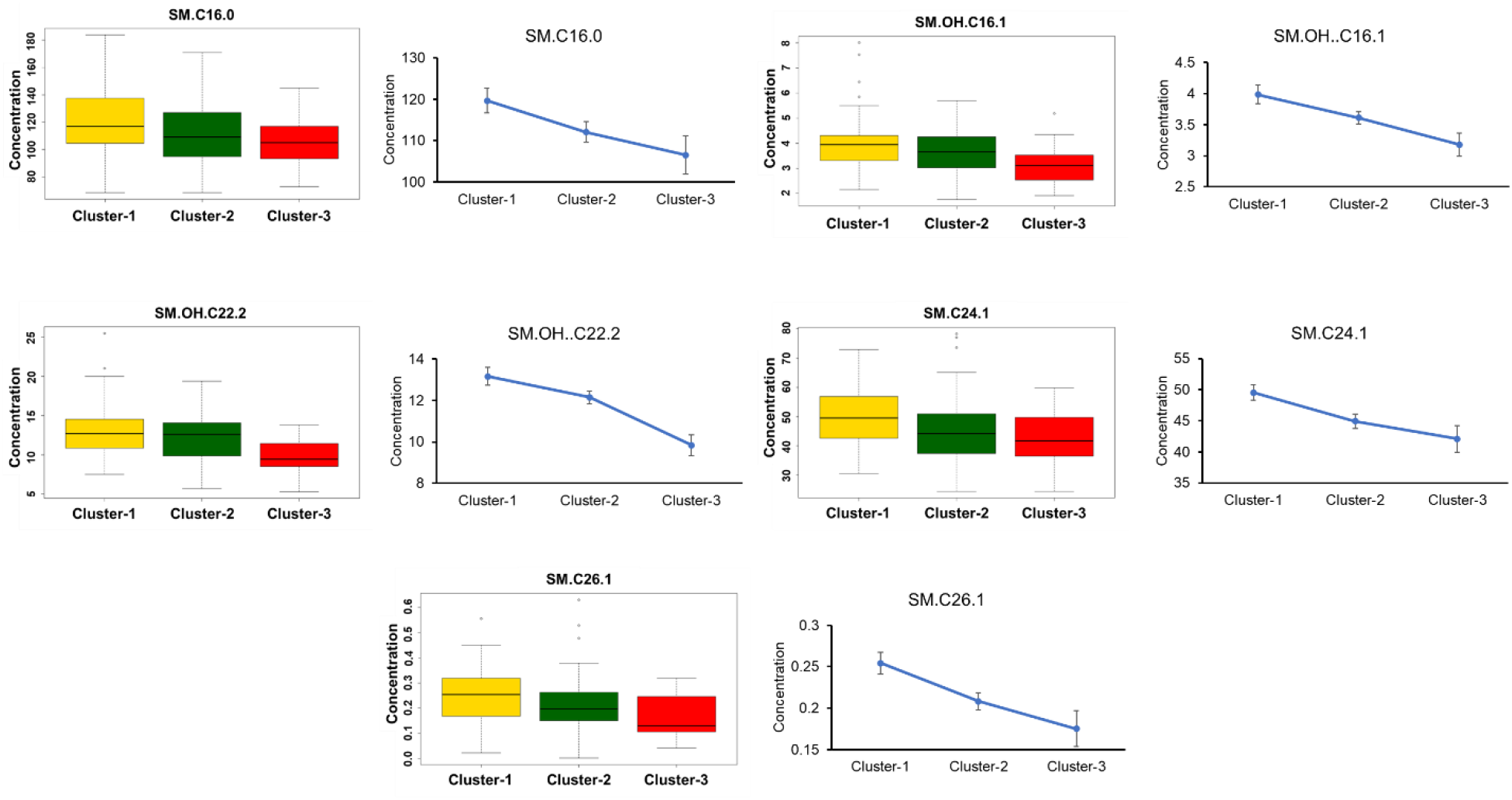
The significantly altered sphingolipid species between clusters. A box plot and adjacent line plot were used here to display the variations between the cluster for each significant sphingolipid.

**Figure 5:**
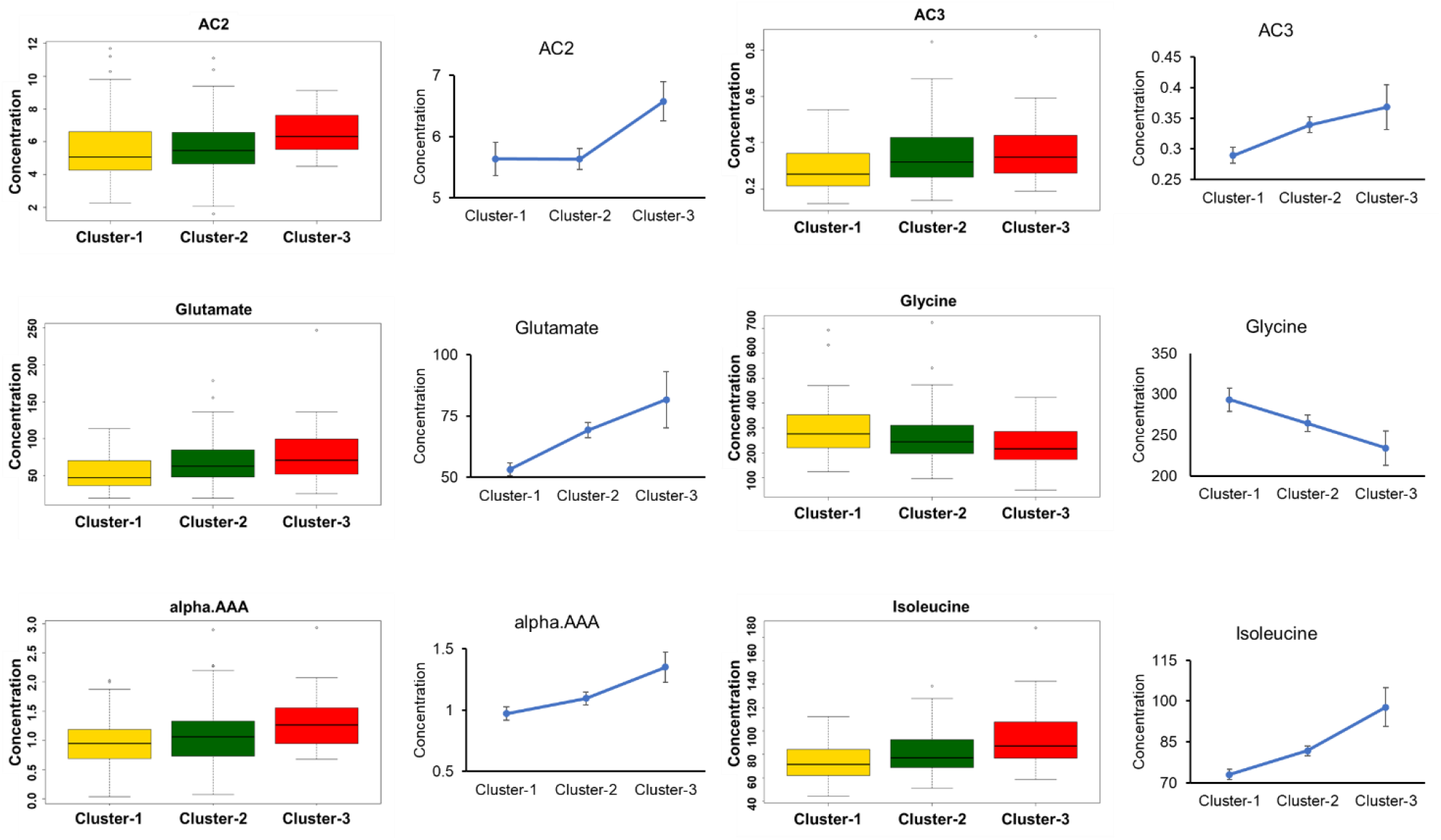
The significantly altered carnitines and amino acids between clusters. A box plot and adjacent line plot were used here to display the variations between the cluster for each significant amino acid.

## Discussion

We have applied a soft-clustering algorithm on baseline (i.e., 6-9 weeks of postpartum) HOMA-B and HOMA-IR values of 225 incident T2D cases (i.e., 226 study participants developed incident T2D within 10 years after GDM pregnancy) among the 1,010 SWIFT study participants without diabetes at baseline, to identify phenotypes of metabolic clusters preceding subsequent progression to T2D. We have chosen these two research variables and selected subsequent incident T2D cases to contrast the pancreatic beta-cell dysfunction and insulin resistance profiles during the early postpartum period [24-27]. We identified three distinguishable clusters within the SWIFT research cohort subsequent incident T2D cases (**Figure-2A)**. These are the severe pancreatic beta-cell dysfunction group (cluster-1), the severe insulin resistance group (cluster-3), and the mixed group (cluster-3). This observation was aligned with previous clustering studies where a hard clustering rule was applied to GDM cohort and identified similar three clusters [17,18]. Altogether, it suggests that GDM and GDM to T2D progression exhibit a similar kind of disease heterogeneity. The clusters identified in the SWIFT cohort are also clinically distinguishable at their 6-9 weeks postpartum based on other glucometric research parameters (i.e., fasting plasma glucose, fasting insulin, and 2-hr glucose and insulin from the OGTT) (**Figure-2D**), fasting lipid profile (i.e., Triglycerides, VLDL-cholesterol, HDL-cholesterol), fasting plasma adipokines (Adiponectin and Leptin) (**Figure-2E and F**), and anthropometric measurements (i.e., weight, BMI) and age (**Table-1**). Among them, cluster-2 (i.e., intermediate form of pancreatic beta-cell dysfunction and insulin resistance, also called mixed group) is the dominant cluster with 120 cases (∼53% cases), followed by cluster-1 (i.e., severe form of pancreatic beta-cell dysfunction, 36% cases) and cluster-3 (i.e., severe form of insulin resistance, ∼11% cases). Therefore, the proportion of incident mixed metabolic cases (i.e., cluster-2) is the highest in the SWIFT Study cohort, followed by cluster-1 (i.e., severe pancreatic beta-cell dysfunction) and case cluster-3 (i.e., severe insulin resistance).

Blood plasma targeted metabolomics data has been used to understand the etiology of the early stages of T2D pathophysiology. Since this analysis did not include the 784 women with GDM in the SWIFT prospective cohort who did not progress to T2D during the same follow up time period (i.e., non-T2D), the interpretations of these molecular mechanisms are focused solely on the comparisons between the case clusters. Additionally, since our data is generated from the baseline fasting plasma of incident T2D cases who had 2-h 75 g OGTTs to confirm no diabetes diagnosis at baseline, this specific analysis of only those who later were diagnosed with T2D by research OGTTs and/or clinical diagnoses is targeting the early stage T2D pathophysiological comparisons. As such, higher concentrations of a molecule at the early stage of T2D pathophysiology between three case clusters indicates its higher importance in either protection against T2D progression or causation of T2D progression. The nature of importance of a molecule depends on its known roles in T2D pathophysiology and relationship to lifestyle behaviors and other factors related to incident T2D, such as lactation intensity and duration (lower risk T2D), maternal obesity, and lipid metabolic profiles related to GDM severity (higher risk of T2D).

It is well-known that insulin resistance correlates with decreased diacyl phosphatidylcholines, a type of glycerophospholipid [28], suggesting a protective role against insulin insensitivity. We have found that several diacyl phosphatidylcholines/plasmalogens (i.e., PC aa C32:1, PC aa C34:3, PC aa C38:3) are higher in severe insulin resistance cluster (i.e., cluster-3) than those of severe pancreatic beta-cell dysfunction cluster (i.e., cluster-1) (**Figure 6**). Our findings imply that these PCs play an important protective role against insulin resistance at the early stage of cluster-3. On the other hand, it is also known that acyl-alkyl phosphatidylcholines (i.e., plasmalogens) and lysophosphatidylcholines (i.e., degraded product of either diacyl phosphatidylcholines or plasmalogens) can stimulate insulin secretion by targeting G protein-coupled receptors [29-31], indicating their protective role against pancreatic beta cell dysfunction. Notably, we have found that several acyl-alkyl phosphatidylcholines (i.e., PC ae C30:0, PC ae C38:0, and PC ae C42:3), and one lysophosphatidylcholine (i.e., lysoPC a 28:1)] are higher in severe pancreatic beta-cell dysfunction cluster (i.e., cluster-1) than those of severe insulin resistance cluster (i.e., cluster-3) (**Figure 6**). It indicates that these types of glycerophospholipids could play an important protective role against pancreatic beta-cell dysfunction of cluster-1 at its early stage long before the onset of T2D, with cluster-1 characteristic of Asians with GDM and lower average pre-pregnancy BMI (**Table 1**).

**Figure 6:**
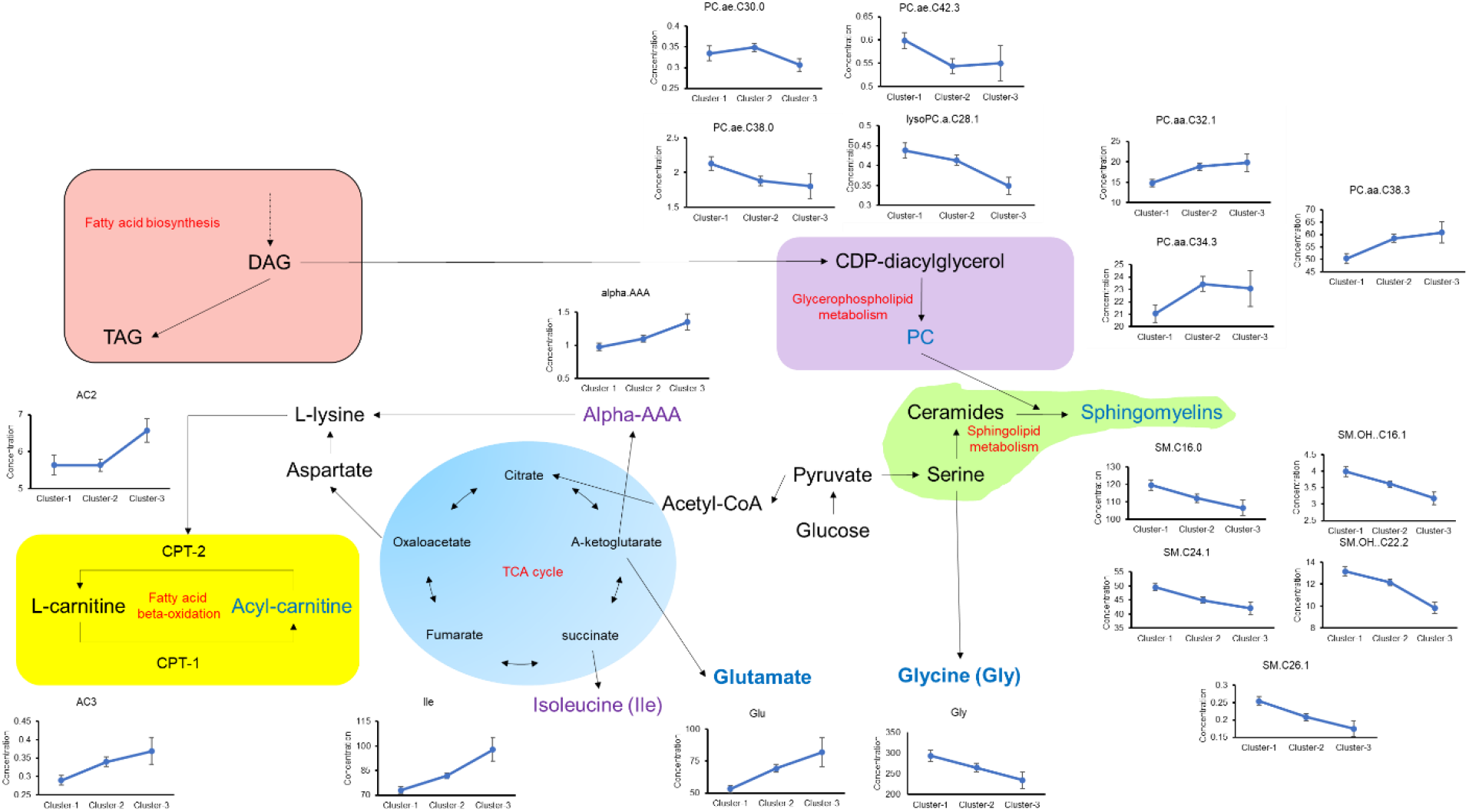
A metabolism roadmap for these three clusters with significantly altered species. The yellow block represents the carnitine metabolism, the blue block represents the TCA cycle, the green block represents the sphingolipid metabolism, the purple block represents the phospholipid metabolism, and the red block represents the fatty acid metabolism. The amino acids in blue colour represent non-essential (i.e., the body can produce) amino acids. The amino acids in purple colour represent essential (i.e., food-derived) amino acids.

In the case of sphingolipids, we have studied only sphingomyelins whose biosynthesis rely on corresponding ceramides and the action of specific ceramide synthase (i.e., CERS1, CERS2, CERS3, CERS4, CERS5, and CERS6) and sphingomyelin synthases (i.e., SM1 and SM2 depends on the biosynthesis sites). For instance, SM C24:1 is synthesized from ceramide C24:1 by the action of ceramide synthase 2 and sphingomyelin synthases. As such, the concentration of sphingomyelins is associated with the biosynthesis of ceramides. The deficiency of ceramide biosynthesis is well-known to cause pancreatic beta cell dysfunction [12, 13]. We have found that five sphingomyelins (i.e., SM C16:0, SM(OH)C16:1, SM(OH)C22:2, SMC24:1, and SMC26:1) are significantly higher in case cluster-1 (i.e., severe pancreatic beta cell dysfunction) than those of case cluster-2 and -3 in this study (**Figure 6**). All of them exhibit concentration-wise similar trend (i.e., case cluster-1 > cluster-2 > cluster-3). It indicates that sphingolipids could have a protective role against pancreatic beta-cell dysfunction. Again, CERS2 is responsible for the biosynthesis of C20-C26 sphingolipids (i.e., CER C20 – C26, SM C20 – C26, etc.), whereas CERS5 and CERS6 are responsible for C16 sphingolipids (i.e., CER C16, SM C16, etc.). Additionally, it is well evidenced that these CERS2 is co-expressed with CERS5 and CERS6 [32], and associated with T2D pathophysiology[12, 33, 34].

Glycerophospholipids (i.e., PC and lysoPC in this study) together with sphingolipids (i.e., sphingomyelins) are integral components of biological membranes and play vital roles in signal transduction and nutrient transportation. PC is synthesized in liver as a component of VLDL and HDL. After being secreted into blood from liver as a part of lipoproteins, PC is degraded to LPC (i.e., lysoPC) via the action of phospholipase A2 (PLA2). LPC upregulates cholesterol biosynthesis[35], downregulates fatty acid oxidation[36], enhances mitochondrial dysfunction[37], and activate inflammatory signaling cascades[38]. As such, higher LPC has known association with diabetes and cardiovascular diseases[39]. On the other hand, sphingomyelin consists of a ceramide and the head group of PC / PE. Prior work has shown a protective role of sphingolipids against pancreatic beta-cell dysfunction and T2D risk [12, 34].

Our study also exhibited that severe insulin resistance among incident T2D cases (i.e., cluster-3) was associated with higher concentration of some acyl-carnitines (i.e., AC2 and AC3) compared to severe pancreatic beta cell dysfunction for incident T2D cases (i.e., cluster-1) (**Figure 6**). The acyl-carnitines are the transformed form of carnitines to carry long-chain fatty acids into the mitochondria for fatty acid beta-oxidation, a protective mechanism against insulin resistance [40]. As such, the higher concentrations of acyl-carnitines in severe insulin resistance incident T2D cases could indicate their protective role at the early stage of disease pathophysiology. This study has also identified four significantly altered amino acids between case clusters. Among them, three amino acids (i.e., isoleucine, alpha-AAA, and glutamate) are significantly higher in cluster-3 than those of cluster-1 (**Figure 6**). Isoleucine is known for its protective role against insulin resistance by stimulating insulin-independent glucose uptake in skeletal muscle cells [41]. Glutamate is also known for its capacity of reducing fasting blood sugar, postprandial blood sugar, and triglycerides [42]. On the other hand, alpha-AAA, an oxidation product of lysine, is a predictive biomarker for insulin resistance in obesity and may appear in the circulation from age-related tissue degradation [43, 44, 45]. Mechanistically, it is a part of carbonyl stress pathway for diabetes pathogenesis [46]. As such, alpha-AAA does not play any protective role in severe insulin resistance case cluster (i.e., case cluster-3). It is an aging marker of insulin resistance for T2D progression. Another amino acid (i.e., glycine) is significantly lower in case cluster-3 than those of case cluster-1 (**Figure 6**). Glycine plays a vital role in ameliorating oxidative stress-induced pancreatic beta cell dysfunction by enhancing plasma antioxidants (i.e., glutathione, catalase, and superoxide dismutase) [47]. Additionally, it improves pancreatic beta cell mitochondrial degeneration and insulin granule degranulation process [47], which decreases with aging [48]. As such, higher concentration of glycine in case cluster-1 is protective against oxidative stress aging-related pancreatic beta cell dysfunction.

In conclusion, this study identifies three mechanisms for incident T2D cases, that could vary in the stage along the pathway of the onset of T2D. We also identify postnatal blood test parameters under research conditions (fasting insulin, glucose and lipids) distinguishing the three clusters that may be measured by clinical laboratories. As far as the mechanistic point-of-view, pancreatic beta cell function requires sphingolipids, acyl-alkyl phosphatidylcholines/plasmalogens, lysophosphatidylcholines, and glycine. A higher demand of these biomolecules at the early stage of disease (i.e., GDM confirmed by research 2-h 75 g OGTTs with No T2D at study baseline 6-9 weeks post-delivery) suggests their compensating functions which is an indicator for pancreatic beta cell dysfunction related to future T2D progression. In contrast, diacyl phosphatidylcholines, acyl-carnitines, isoleucine and glutamate are critical for insulin sensitivity. A higher demand of these biomolecules at the early stage of disease (i.e., GDM with No-T2D) suggests their compensating functions which is an indicator for insulin resistance related to future T2D progression. Notably, all these biomolecules (i.e., sphingolipids, glycerophospholipids, acyl-carnitines, and amino acids) are found in the cluster-2 (i.e., mixed form of both phenomena in intermediate levels) in the mediocre demands, indicating a true-nature of a mixed group. This information will aid in adopting proper interventions using precision medicine approach.

### METHODS

We utilized 225 incident T2D cases identified during the 10-12 year follow up in the prospective SWIFT research cohort of women with GDM, and applied machine learning-based clustering algorithm and statistical analyses. The SWIFT cohort follows a prospective study design and protocol in terms of its design, recruitment, selection criteria, and serial in-person research examinations with 2-h 75 g OGTTs and research protocol for anthropometry, and comprehensive medical, social and lifestyle behavioral assessments, described elsewhere [19, 49]. The details of clustering analyses and their characterization have been described here.

### SWIFT cohort

1,035 racially and ethnically diverse (35% Asian, 32% Hispanic, 24% White, 8% African, and 2% others) GDM women (aged 20-45 years) were enrolled in the SWIFT research cohort. GDM was diagnosed using Carpenter and Coustan criteria on the 3-hour 100-g OGTT test at the last trimester of pregnancy. All of them delivered a singleton, full-term (after 35 weeks of gestation), live-born infant at the Kaiser Permanente Northern California (KPNC) hospital system (i.e., 13 KPNC medical center/office facilities in a 5,000-square-mile KPNC region) during the period of 2008-2011. Overall, 32.8% were primiparous 36.8% were biparous and 30.4% were multiparous. The eligibility criteria for recruitment of women into the early postpartum (study baseline 6-9 weeks postpartum) research cohort included ‘no diabetes history’, ‘no other serious health conditions’, ‘not receiving glucose tolerance altering medication’, and ‘no plan for another pregnancy’ and intention to exclusively or mostly breastfeed for at least 4 months or mostly formula feed from 1 month post-delivery. Research staff contacted women during pregnancy and again at 4 weeks post-delivery to determine their eligibility for the study. Participants provided written informed consent for 3 in-person research visits which included a 2-h 75 g research OGTT and other assessments per research protocol which occurred at baseline and annually for two years post-baseline. The SWIFT Study participants continued in ongoing follow up after baseline for clinical diagnosis of new onset T2D via the KPNC electronic health records (ICD 9/10 codes, prescription medications, and laboratory clinical tests for glucose tolerance) more than 10 years post-baseline. The in-person research visits occurred at 6-9 weeks of postpartum (study baseline), and at 1 year and 2 years post-baseline. These in-person research exams included a research 2-hour 75-g OGTT, and anthropometric measurements (i.e., body weight, height, adiposity and waist circumference) under research protocols conducted by trained research staff using calibrated research instruments, and interviewer- and self-administered surveys that queried participants on socio-demographics, medical and reproductive histories, family history of diabetes, sleep habits, depression (CES-D), lactation and infant feeding, and lifestyle behaviors (diet and physical activity). At each in-person research visit, EDTA-treated plasma and buffy coat cells were collected during the research 2-hour 75-g OGTTs and analyzed within several weeks by the Northwest Lipid Research Laboratories at the University of Washington, Seattle, WA, with research quality assays for plasma glucose, insulin, lipids, adipokines and other biomarkers from blood collected under stringent research protocols (fasting 10 hours or more, and fasting blood draw before 10 a.m.). The aliquots of these plasma samples were stored in the SWIFT biobank (−80°C) for other biochemical analyses (i.e., adipokines, lipids, metabolomics and lipidomics). The SWIFT research cohort is embedded within the Kaiser Permanent Northern California (KPNC) healthcare system, and clinical diagnoses of diabetes supplemented the research OGTTs, and identified new onset diabetes in the cohort beyond the two years post-baseline through 2020 for this analysis. The American Diabetes Association (ADA) criteria for diagnosis of diabetes were applied to classify participants at each research visit and during later follow up for additional clinical diagnoses of new onset T2D. The study design and all procedures were approved by the KPNC Institutional Review Board, Oakland, CA, USA, and the subsequent assays of stored specimens were also approved by the Office of Research Ethics at University of Toronto. All the participants provided written consent for the SWIFT study (KPNC, Oakland, CA, USA) data collection and follow up, and for use of research data and stored specimens in subsequent research studies[19].

### Study Design

This sub-analysis included 225 of 226 women from the SWIFT cohort who developed incident T2D during ∼10 years of prospective cohort follow-up. At study baseline (6 to 9 weeks postpartum), we excluded 23 women: 21 women classified with T2D via the 2-h 75 g OGTT, and 2 women who were drop-outs at baseline. Blood plasma (EDTA treated fasting and 2-h) was available for 1010 women without diabetes at baseline, and of these, 990 had follow up testing of glucose tolerance via the SWIFT annual research 2-h 75 g OGTTs for two years, and/or subsequent KPNC clinical laboratory testing obtained from the electronic health records through October 21, 2020 [20]. Among 990 with post-baseline testing during follow up, 226 women progressed to T2D within ∼10 years post-baseline. The remaining 784 women did not develop T2D (No T2D) during the same follow up time period. For this prospective longitudinal study, women who progressed to new onset T2D were classified as an incident T2D case, whereas women who did not develop diabetes were No T2D or “control” in this context. One incident T2D case was excluded due to missing insulin measurements; leaving 225 women with GDM for this analysis who later developed incident T2D. These 225 incident T2D cases were stratified for identification of T2D heterogeneity using two research variables measured at study baseline, fasting glucose and insulin, to calculate measures of pancreatic beta-cell function (i.e., HOMA-B) and insulin resistance (i.e., HOMA-IR)[19, 20]. The identified clusters using case participants is termed as incident “clusters”.

### Metabolomics

Metabolomics data was available on a subset of 173 T2D cases. This metabolomics panel was designed based on literature review of previous T2D metabolic studies. The most of these selected metabolites are covered by the AbsoluteIDQ p180 plate, consisting of a total of 188 metabolites using mass-spectrometry-based detection on the supplied 96-well extraction plate according to manufacturer instructions (Biocrates Life Sciences, Innsbruck, Austria). It requires 10 μl of each serum sample with 10 μl of the supplied internal standard solution, QC samples, and calibration standards. The data was collected through multiple reaction monitoring transitions for each analyte and internal standard using a scheduled retention time window as per AbsoluteIDQ p180 Kit. These collected data was used to calculate the concentration using Biocrates software MetIDQ. In the case of amino acids (AAs) and hexoses, chromatographic separation was carried out through an Agilent 1290 HPLC (an Agilent Eclipse XDB-C18 100 × 3.0 mm, 3.5 μm column) using either deuterated or 13C stable isotope-labeled internal standard of the exact analyte or a closely eluting compound of similar class, followed by measurement with a SCIEX QTRAP 5500 mass spectrometer. Besides, a flow injection analysis tandem mass spectrometry (FIA-MS/MS) were deployed for acylcarnitines (40), sphingolipids (15), and glycerophospholipids (90). All assays were performed and assessed, without disclosure of group allocation, by the Analytical Facility for Bioactive Molecules (The Hospital for Sick Children, Toronto, ON, Canada). We corrected *P*-value for multiple testing using Benjamini–Hochberg method and used FDR cut-off < 0.05.

### Incident T2D Case heterogeneity identification

The incident T2D case heterogeneity among the clusters was identified using a gaussian finite mixture modelling algorithm, named mclust (a popular R package for model-based clustering), for two baseline (6-9 weeks postpartum) indices of glucose homeostasis – HOMA-IR and HOMA-B on incident T2D of the entire SWIFT cohort (i.e., 225 cases with complete glucose and insulin). This mclust() function constructs different models (i.e., EII, EVV, EEI, VII, VVI, VEE, EVE, VVE, VEI) from the data matrix by mixing components and the covariance parameterisation using the Bayesian Information Criterion (BIC), which is a penalised forms of the log-likelihood. Typically, the likelihood increases with the addition of more components. The penalised forms of the log-likelihood are calculated as a penalty term for the number of estimated parameters subtracted from the the log-likelihood. These models are arranged in negative value of BIC. The lower the negative value that indicates lesser penalty is the better model. The number of components (or clusters) are selected from the top models and their number of components before plateauing of their BIC values. The details of mclust can be found in elsewhere [50].

### Statistical analysis

Where relevant, we applied nonparametric t-test and ANOVA on research variables for characterization of all case clusters. The significance of metabolomics data is corrected with multiple testing using Benjamini–Hochberg method and used FDR cut-off < 0.05.

## Data Availability

Requests to access the dataset from qualified researchers may be sent to Dr. Erica P. Gunderson, Principal Investigator, at the
Division of Research.

## ACKNOWLEDGMENTS

This study is supported by research grants from Canadian Institutes of Health Research (CIHR): FRN 143219 (M.B.W.), https://www.cihr-irsc.gc.ca. The National Institute of Child Health and Human Development (NICHD): R01HD050625 (E.P.G.), https://www.nichd.nih.gov/. The National Institute of Digestive, Diabetes and Kidney Disease (NIDDK): R01DK118409 (E.P.G.), https://www.niddk.nih.gov/. BJC is supported by a Tier II Canada Research Chair. The funders had no role in study design, data collection, and interpretation, or the decision to submit the work for publication.

## AUTHOR CONTRIBUTIONS

SRK and EPG conceptualized and designed the research work executed by SRK. SRK wrote the manuscript, edited by all. All authors reviewed the manuscript and gave final approval of the version to be published.

## STUDY APPROVALS

The study participants provided written informed consent for all study procedures, including use of biospecimens and electronic health records (EHR) data, and data analysis for future studies. The Kaiser Permanente Northern California (KPNC) Institutional Review Board approved the Study of Women, Infant Feeding and Type 2 Diabetes after Gestational Diabetes (SWIFT) research protocol and procedures, and this data analysis. This study followed the Strengthening the Reporting of Observational Studies in Epidemiology (STROBE) guidelines.

## TOP GUIDELINES

Requests to access the dataset from qualified researchers trained in human subjects’ confidentiality protocols may be sent to Dr. Erica P. Gunderson, Principal Investigator, at the Division of Research. The patient data is owned by the Kaiser Foundation Health Plan, Inc., Kaiser Foundation Hospitals, Inc., and The Permanente Medical Group, Inc. Because of their third-party rights, it is not possible to make the data publicly available without restriction.

